# Navigating Fairness in AI-based Prediction Models: Theoretical Constructs and Practical Applications

**DOI:** 10.1101/2025.03.24.25324500

**Authors:** S.L. van der Meijden, Y. Wang, M. S. Arbous, B. F. Geerts, E.W. Steyerberg, T. Hernandez-Boussard

## Abstract

Artificial Intelligence (AI)-based prediction models, including risk scoring systems and decision support systems, are increasingly adopted in healthcare. Addressing AI fairness is essential to fighting health disparities and achieving equitable performance and patient outcomes. Numerous and conflicting definitions of fairness complicate this effort. This paper aims to structure the transition of AI fairness from theory to practical application with appropriate fairness metrics. For 27 definitions of fairness identified in the recent literature, we assess the relation with the model’s intended use, type of decision influenced and ethical principles of distributive justice. We advocate that due to limitations in some notions of fairness, clinical utility, performance-based metrics (area under the receiver operating characteristic curve), calibration, and statistical parity are the most relevant group-based metrics for medical applications. Through two use cases, we demonstrate that different metrics may be applicable depending on the intended use and ethical framework. Our approach provides a foundation for AI developers and assessors by assessing model fairness and the impact of bias mitigation strategies, hence promoting more equitable AI-based implementations.

## 1. Introduction

With the rise of Artificial Intelligence (AI)-based prediction models in clinical settings, concerns about bias and fairness have become critical. Biases from data sources, collection procedures, algorithm design, and decision-making can lead to inequitable health outcomes for marginalised groups [1]. While these models can reduce health disparities [2], they can also exacerbate them, as demonstrated by significant racial bias in a widely used health management algorithm [1]. This risk of discrimination underscores the need for fair AI, prompting increased attention from regulatory bodies [3, 4].

Although AI fairness and bias mitigation strategies are well-studied, consensus on what constitutes fairness remains elusive [5]. Fair AI, unlike bias, considers the ethical implications of use in marginalised populations. Evaluating fairness is challenging due to the variety of notions, metrics, and frameworks available, which often conflict, requiring trade-offs [8-12]. Studies suggest addressing bias and fairness throughout all phases of model development and deployment [10, 13].

Despite guidance on fairness metrics in various contexts [12], a gap remains in how to choose appropriate metrics for evaluating AI-based prediction models. This, along with the context-dependent nature of fairness and differing perspectives between AI developers and healthcare providers [7, 14], has hindered the operationalisation of AI fairness. Specific strategies, such as excluding protected attributes like race, require careful consideration, as inclusion may enhance model accuracy but risk promoting race-based medicine [15-17].

Our aim was to provide a structured approach to identifying appropriate fairness evaluation methods for AI-based prediction models. We first review the literature on fairness notions and metrics. Then, we propose a framework to select fairness metrics based on the model’s intended use, decision type, and ethical principles of distributive justice. Finally, we apply the framework to two use cases, illustrating the need for ethical trade-offs in fairness evaluation. As the field of AI fairness struggles with numerous definitions and terminology, the important concepts for this paper are outlined in Box 1.

## 2. Evaluation of model fairness and bias mitigation

Overall challenges concerning model fairness can be divided into data, and modelling and evaluation challenges (Figure 1). Bias mitigation strategies should be considered throughout the AI-based prediction model development lifecycle—from problem formulation to monitoring and deployment—to enhance model fairness. We propose implementing checkpoints at each phase (see Supplementary Materials, Figure S1). Bias mitigation strategies are typically categorised as pre-processing (e.g., resampling), in-processing (e.g., weighted learning), and post-processing (e.g., adapting classification thresholds) [19]. Although this paper does not focus on specific strategies, extensive overviews are available [14, 20, 21]. It is crucial to consider how fairness notions relate to these strategies. For instance, weighted learning might achieve equal predictive performance across patient groups but could lead to a general decline in performance, reflecting the ‘fairness-accuracy trade-off’. The clinical, ethical, and societal contexts influence decisions on whether a model is ‘fair enough’ for clinical use, where cut-offs are context-dependent.

**Figure 1.**
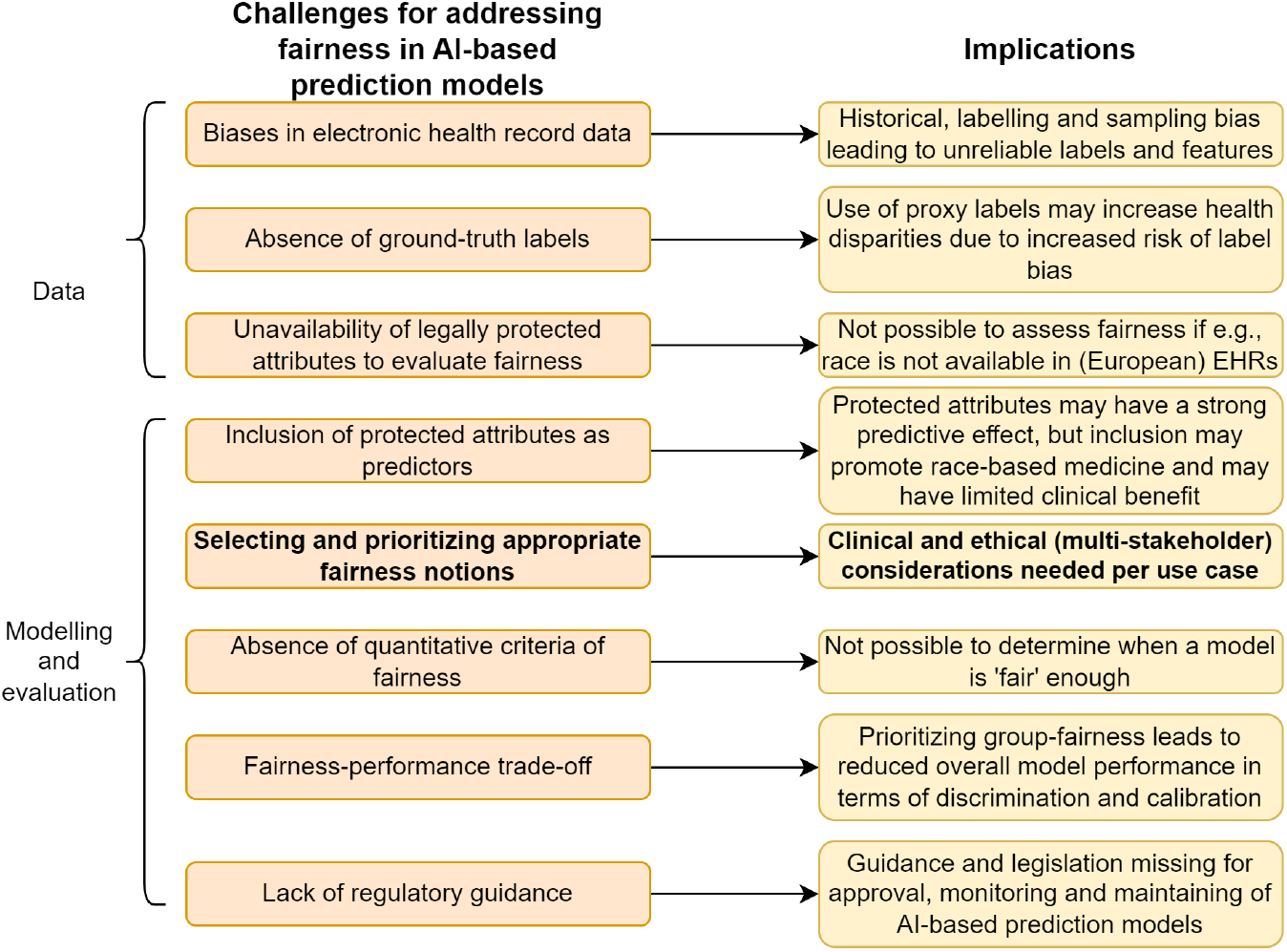
Overall challenges and corresponding implications in addressing fairness for AI-based prediction models. In bold, the focus of this paper is indicated. EHR = Electronic Healthcare Systems.

### Box 1

Main definitions of terms concerning bias and fairness of AI-based prediction models

| Definition | Explanation in the context of this paper |
| --- | --- |
| **AI-based prediction models** | Artificial Intelligence-based prediction models aimed at predicting patient outcomes. Can be either used as clinical decision support systems to classify patients to support a specific decision, or to inform decision-making by providing a risk score between 0-100%. |
| **Bias mitigation** | Strategies that can be applied throughout model development and deployment to achieve a higher level of model fairness. Bias mitigations may be categorised as pre-processing (e.g., data acquisition, resampling), in-processing (e.g., weighted learning), and post-processing (e.g., adapting classification thresholds). |
| **Clinical utility** | Clinical utility refers to the value of a prediction model in a healthcare setting, measured by its benefit to patient outcomes when used in decision-making processes. Clinical utility may be evaluated through decision curve analysis, which assesses the trade-offs between the benefits and harms of basing decisions on predictions from the model. |
| **Equalized odds** | A commonly used notion of fairness requires that the true positive rate (sensitivity) and the false positive rate (1-specificity) are the same for each group. |
| **Fairness** | There is no consensus on the definition of AI fairness (Supplementary Materials, Table S2). In general, the field of fair AI strives to reduce health disparities in marginalised patient groups. |
| **Fairness-accuracy trade-off** | Improving fairness may require adjusting the model to mitigate biases, which can lead to a decrease in overall accuracy. Conversely, optimising for maximum accuracy without considering fairness can result in unjustified inequality in outcomes across certain groups. |
| **Fairness notion** | Refers to a specific formalisation or criterion used to evaluate and ensure fairness (e.g., equalized odds, statistical parity). |
| **Group fairness** | Refers to a category of fairness criteria that aims to ensure that different demographic groups (e.g., based on race, gender, age) have equal performance and are treated equally by the AI-based prediction model. |
| **Individual fairness** | Refers to a category of fairness notions focusing on ensuring that similar individuals receive similar predictions or outcomes from the AI-based prediction models. |
| **Intended use** | Defines the specific clinical purpose the model aims to address, guiding the design, development, validation, and deployment within healthcare settings [18]. |
| **(Non)-polar decisions** | In polar decisions, patients benefit most from having a biased prediction towards high (or low) risk. This is in contrast to the non-polar case, where the interests of the caregiver using the model and patient align such that the patient will benefit most from a model with highest predictive performance. |
| **Protected (or sensitive) attributes** | Specific characteristics or features of individuals legally or ethically safeguarded against discrimination (e.g. race, gender, age). |
| **Protected group** | Patient group defined by a protected attribute. E.g., all Black patients. |
| **Statistical parity** | A commonly used notion of fairness that requires equal proportions of predicted positives (or negatives) across groups. |

## 3. Fairness evaluation methods

We conducted a systematic review to gather fairness notions and evaluation methods for AI-based prediction models, identifying 16 studies (Supplementary Materials Section 2). Definitions of fairness and bias varied significantly, with 27 evaluation perspectives (Supplementary Table S2 and S3), highlighting the need for consistent naming conventions [5].

Fairness notions were often categorised into group-based and individual fairness metrics. Group-based metrics assess outcomes across subgroups, while individual fairness ensures similar predictions or treatments for similar patients [7, 9, 21]. Individual fairness requires similarity metrics, which are complex as patient similarity can be defined in many ways [7, 14, 22]. We focused on group-based notions like sensitivity and specificity across subgroups, relevant for systemic assessments by hospital leadership and policymakers [7]. However, group fairness requires high-quality data on protected attributes like race and ethnicity [15, 16]. There is a tension between individual and group fairness: individual fairness addresses patient-specific needs, while group fairness focuses on demographic equality, potentially missing unique circumstances.

Including protected attributes like race in a model can impact fairness notions differently: improving fairness for some groups while worsening it for others (see Table 1). This inclusion assumes the attribute relates to the outcome in ways other factors cannot explain, such as racial disparities persisting despite accounting for socioeconomic status. Performance metrics used to assess fairness rely on ‘ground truth’, which may be biased due to historical or label biases [24]. Statistical parity, which assesses predicted outcomes regardless of actual outcomes, is the only metric independent of ground truth, but it has limitations.

**Table 1.**
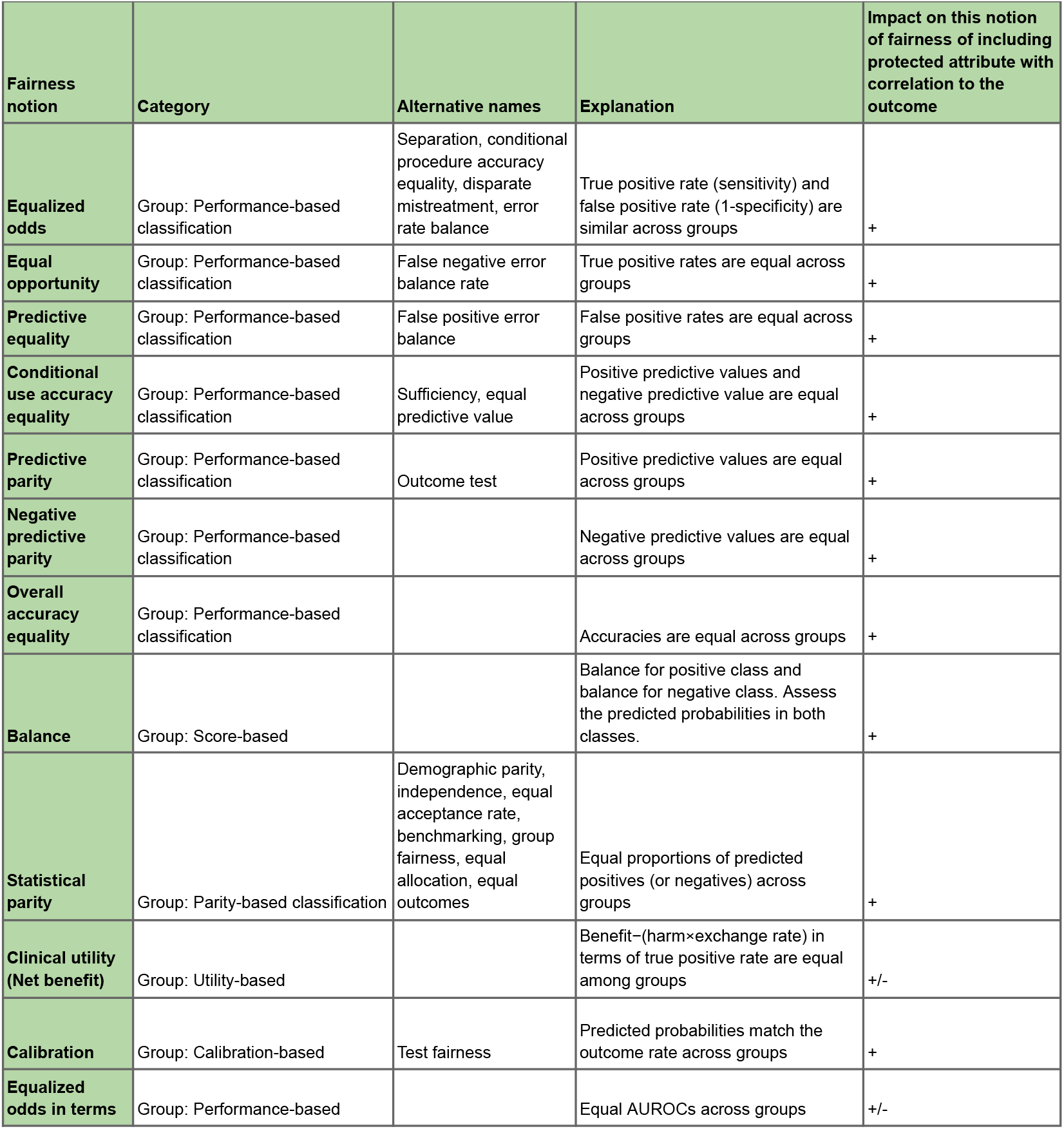
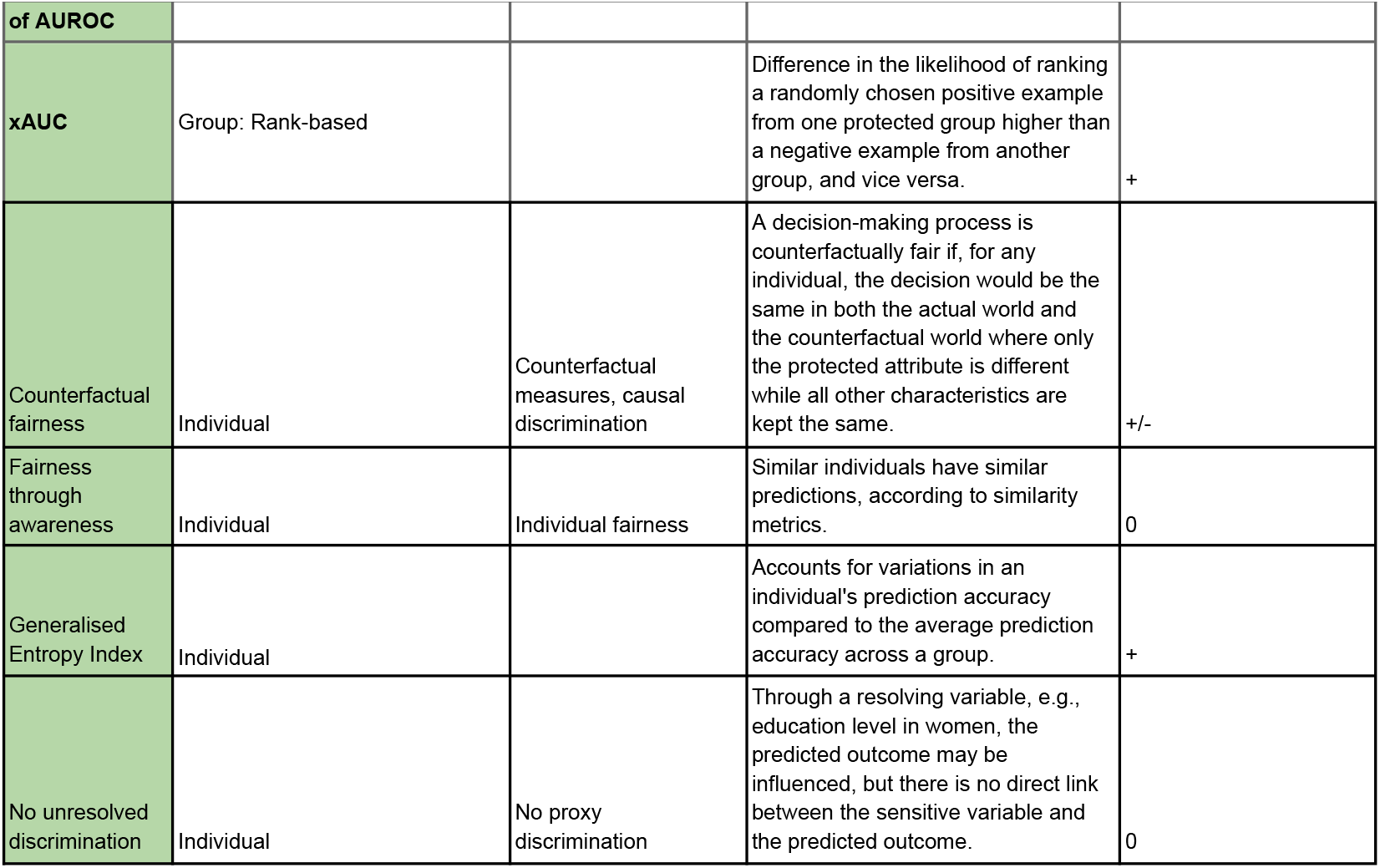
Overview of commonly used group-based and individual notions of fairness. AUROC = area under the receiver operating characteristic curve, FPR = false positive rate, NPV = negative predictive value, PPV = positive predictive value, TPR = true positive rate, xAUC = cross-area under the curve.

## 4. Guidance on prioritising fairness metrics

Per type of use case, it may differ which type of fairness notions are applicable [6, 15, 25]. We provide a simplified framework to determine relevant group-based notions of fairness (Table 2). This framework serves as an initial guide. It is flexible to other notions, such as individual fairness, which may be of interest due to the clinical and ethical context. The choice of fairness notion is important as fairness metrics cannot be all met at once. It is logically impossible for any model to simultaneously satisfy, for example, both the conditions of ‘conditional use accuracy equality’ (positive predictive value and negative predictive values are equal) and ‘equalized odds’ (true positive rates and false positive rates are equal) [2].

**Table 2.**
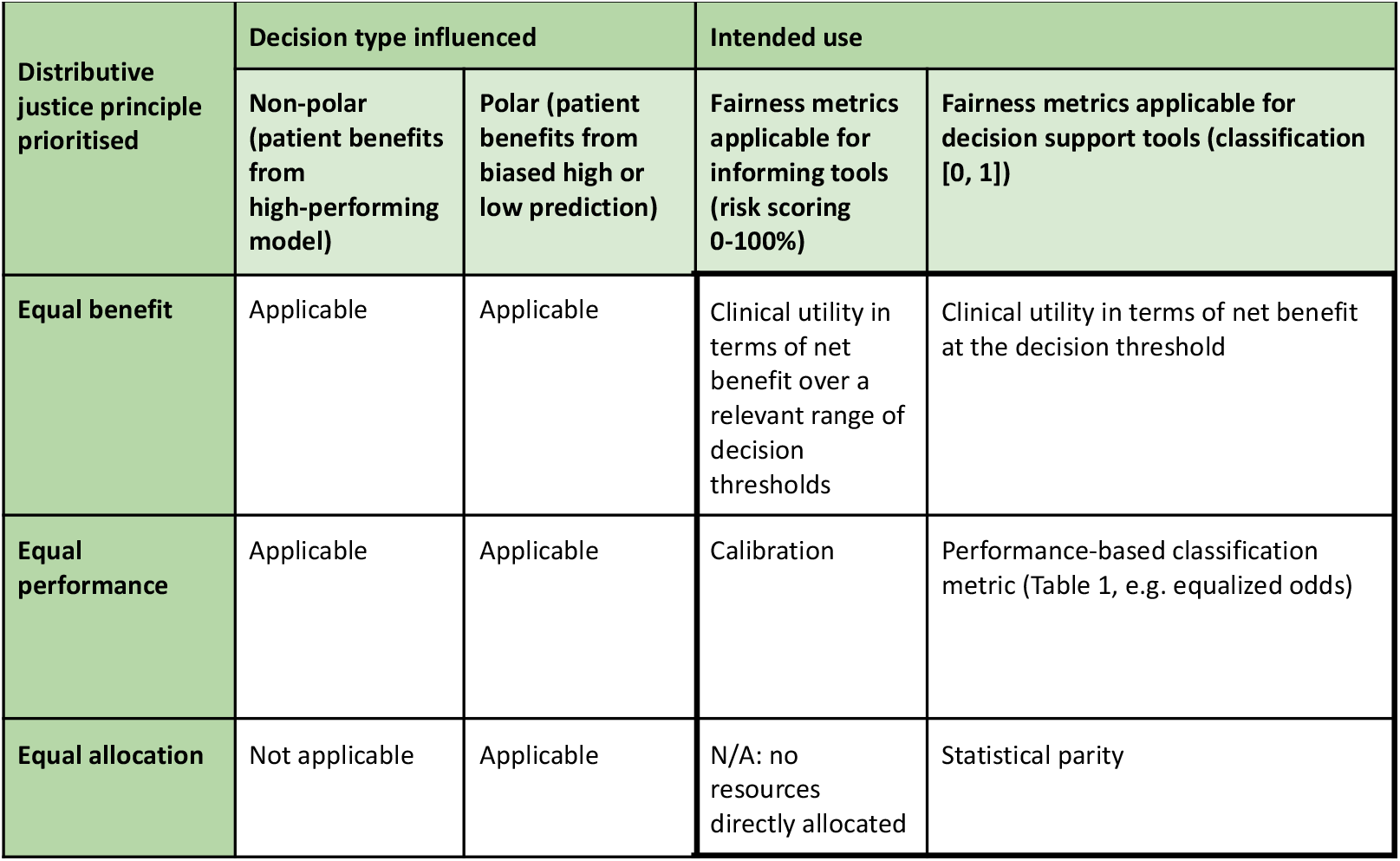
Fairness metrics: Decision diagram to provide guidance and example group notions of fairness to prioritise for different use cases.

| Distributive justice principle prioritised | Decision type influenced |  | Intended use |  |
| --- | --- | --- | --- | --- |
|  | Non-polar (patient benefits from high-performing model) | Polar (patient benefits from biased high or low prediction) | Fairness metrics applicable for informing tools (risk scoring 0-100%) | Fairness metrics applicable for decision support tools (classification [0, 1]) |
| Equal benefit | Applicable | Applicable | Clinical utility in terms of net benefit over a relevant range of decision thresholds | Clinical utility in terms of net benefit at the decision threshold |
| Equal performance | Applicable | Applicable | Calibration | Performance-based classification metric (Table 1, e.g. equalized odds) |
| Equal allocation | Not applicable | Applicable | N/A: no resources directly allocated | Statistical parity |

### 4.1. Intended use: Inform versus decision support

First, the intended use may be ‘informing with a risk score’ or ‘decision support with a classification’. This intended use guides the design, development, validation, and eventual deployment within healthcare settings [18]. For example, a decision support tool may diagnose pneumonia on X-ray images (yes/no) to guide treatment decisions, while an informing prediction tool may predict the risk of developing diabetes (0-100%) based on electronic health record data, without clear action in mind. The key difference is that a decision support tool uses a threshold to assign patients to either a positive or negative classification while informing prediction tools operate without a threshold, making threshold-dependent metrics such as equalized odds and statistical parity inapplicable.

### 4.2. Decision type influenced: Non-polar versus polar

Second, it needs to be determined whether the decision targeted to be influenced is polar or non-polar [25]. In polar cases, patients benefit most from having a biased prediction towards high (or low) risk.

This contrasts with the non-polar case, where the interests of the caregiver and patient are aligned, i.e. provide care as needed by the patient to derive optimal outcomes. In non-polar cases, patients benefit most from a model with highly reliable predictions and high overall performance. For example, in distributing scarce but valuable healthcare resources (polar), striving for statistical parity (equal positive rates) across groups may be desirable. Conversely, in non-polar disease classification to optimise treatment decisions, biased predictions lead to an improper balance between overtreatment and undertreatment. In most clinical applications, the type of decision influence is non-polar [25].

### 4.3. Distributive justice principle prioritised

Third, principles of distributive justice may guide the choice of relevant fairness notions [19]. Equal benefit, derived from equal outcomes between patient groups, can be assessed using net benefit, placing the harms and benefits of the prediction model on the same scale [26]. Clinical utility may be assessed over various decision thresholds or at a specific threshold for decision support tools.

In both non-polar and polar cases, striving for equal benefit may be necessary. The principle of equal performance relates to calibration of predictions provided by informing prediction tools and relates to performance-based metrics such as equalized odds for decision support tools. Additionally, equal allocation of healthcare resources across groups aligns with statistical parity in decision support tools.

## 5. Illustrative use cases

We demonstrate that fairness notions of interest may differ for two AI-based prediction models. The first is to predict postoperative infections based on data from the Leiden University Medical Center, The Netherlands. The second predicts opioid use disorder risk based on the MIMIC-IV data [27]. These use cases were selected to address key considerations in fairness evaluation while focusing on two considerable healthcare challenges. Postoperative infections and opioid use disorder have significant clinical and societal implications, making them highly relevant areas for exploring fairness in predictive modelling. We required complete access to the models’ code and data to conduct a fairness evaluation according to our framework, which goes beyond the scope of prior work we have performed on these use cases [28, 30]. The opioid use disorder model was developed on open-source MIMC-IV data.

Additionally, the postoperative infection model was chosen because it will soon be employed in clinical settings, enabling us to explore fairness in real-world patient impact. Finally, by including this model trained on non-US data, we sought to address areas of bias less dominated by race and ethnicity, providing a broader perspective on fairness in diverse healthcare settings.

We present the quantitative fairness evaluation on an independent test dataset for both XGBoost machine learning models. We evaluated fairness metrics using Python 3‐8 and the Sklearn and Fairlearn packages. 95% confidence intervals were calculated using 1000 bootstrap resampling.

Subgroup characteristics, performance, and fairness metrics are summarised in the Supplementary Materials Table S3 and S4.

### 5.1 PERISCOPE: Predicting postoperative infections

PERISCOPE is an AI-based informative prediction tool that provides a probability of postoperative infection following a surgical procedure in the hospital setting [28]. The ultimate aim is to reduce the severity and impact of infection by supporting clinical actions related to the diagnosis and/or treatment of postoperative bacterial infections, but no classification to guide treatment is directly provided.

Decisions that may follow from using PERISCOPE are non-polar. We hence should focus on the calibration of predictions (to strive for equal performance) and clinical utility (to strive for equal benefit). We evaluate fairness metrics in gender subgroups. As men are known to have higher infection rates, in correspondence with our findings, gender was included as a feature in the model (Table 3). The model showed slight overestimation in female patients, even though gender was included in the prediction model (Figure 2). Clinical utility was positive overall and by gender subgroup (Figure 3). Based on these findings, we determined that there was no need for further bias mitigations for gender subgroups.

**Table 3.**
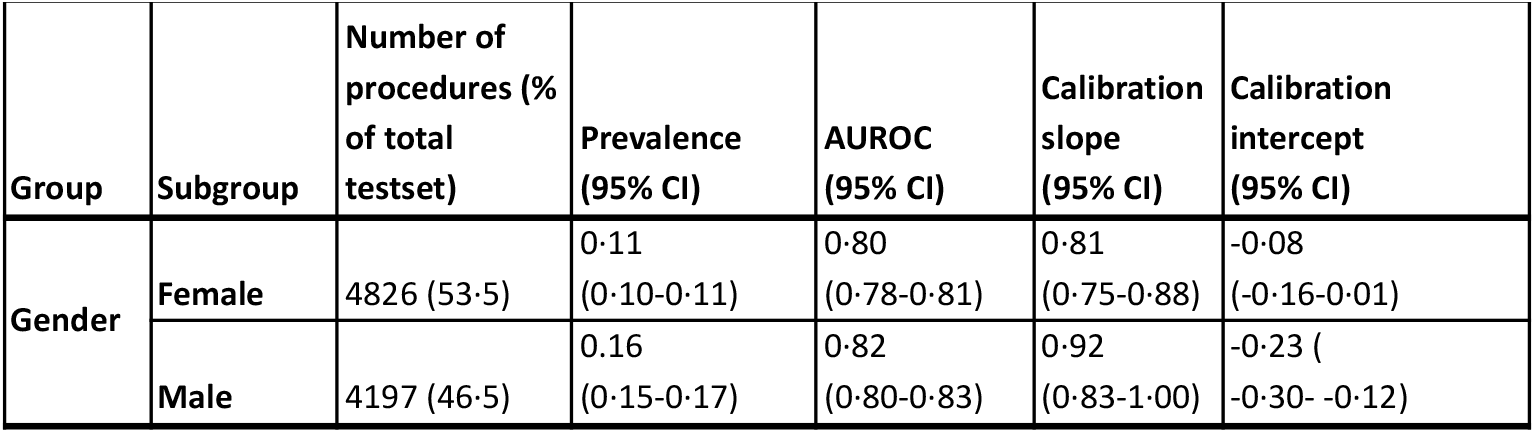
PERISCOPE prevalence and performance metrics for predicting postoperative infections in test dataset. CI = confidence interval.

**Figure 2.**
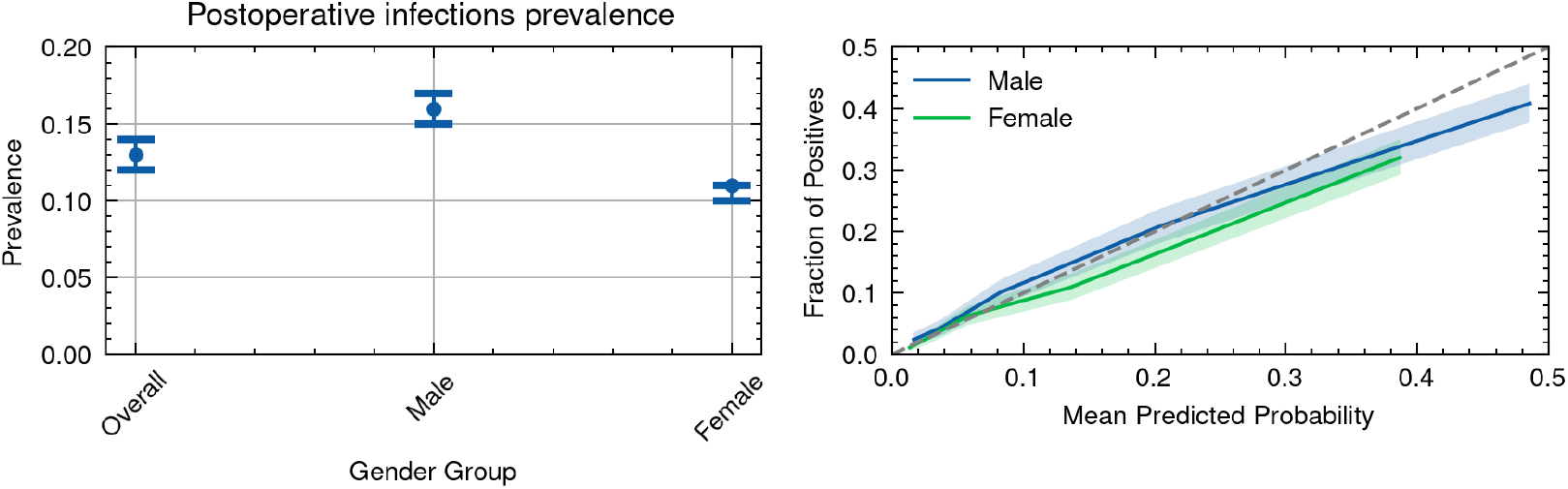
Baseline incidence rates with 95% confidence intervals (left) and calibration curves (right) for PERISCOPE predictions in gender subgroups.

**Figure 3.**
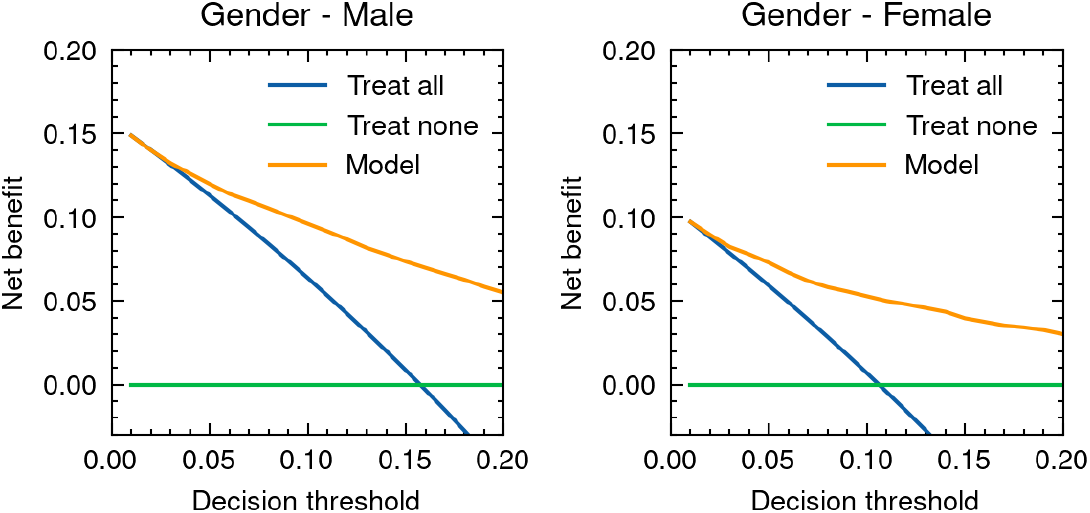
Clinical utility by means of net benefit for PERISCOPE’s postoperative infection prediction in gender subgroups.

### 5.2 Allocating resources for opioid use disorder risk

For opioid use disorder (OUD) risk, previous studies indicated that Black patients were less likely to qualify for additional education, treatments, and care management programs for opioid misuse [29]. The principle of equal allocation was considered desirable in race subgroups for this use case. Details on the patient population, features and feature engineering are available in a previous study [30]. Race was excluded as a predictive feature due to its limited impact on performance and net benefit for predicting OUD risk (Table S5 and Figure S2 in the Supplementary Materials). The AI-based decision support tool’s intended use was to classify patients for receiving additional services, making it a polar decision where patients with high OUD risk benefit from resource allocation to fight health disparities. We assessed statistical parity (positive rate) and clinical utility, aiming to allocate resources equally across race groups with high risk. We observed a slightly lower allocation in the Hispanic group, where baseline incidence was lowest (Table 4, Figure 4). Using a 10% classification threshold, net benefit was calculated (Figure 5) showing zero net benefit for Hispanic patients, likely due to the lower AUROC in this group (Table 4). Based on these findings, bias mitigation for race subgroups may be necessary to enhance model fairness. We explored the effect of statistical oversampling on the training dataset, which improved Net benefit for Hispanic patients, but worsened performance for Black/African American patients (Supplementary Materials Figure S3). Overall AUROC did not change.

**Table 4.**
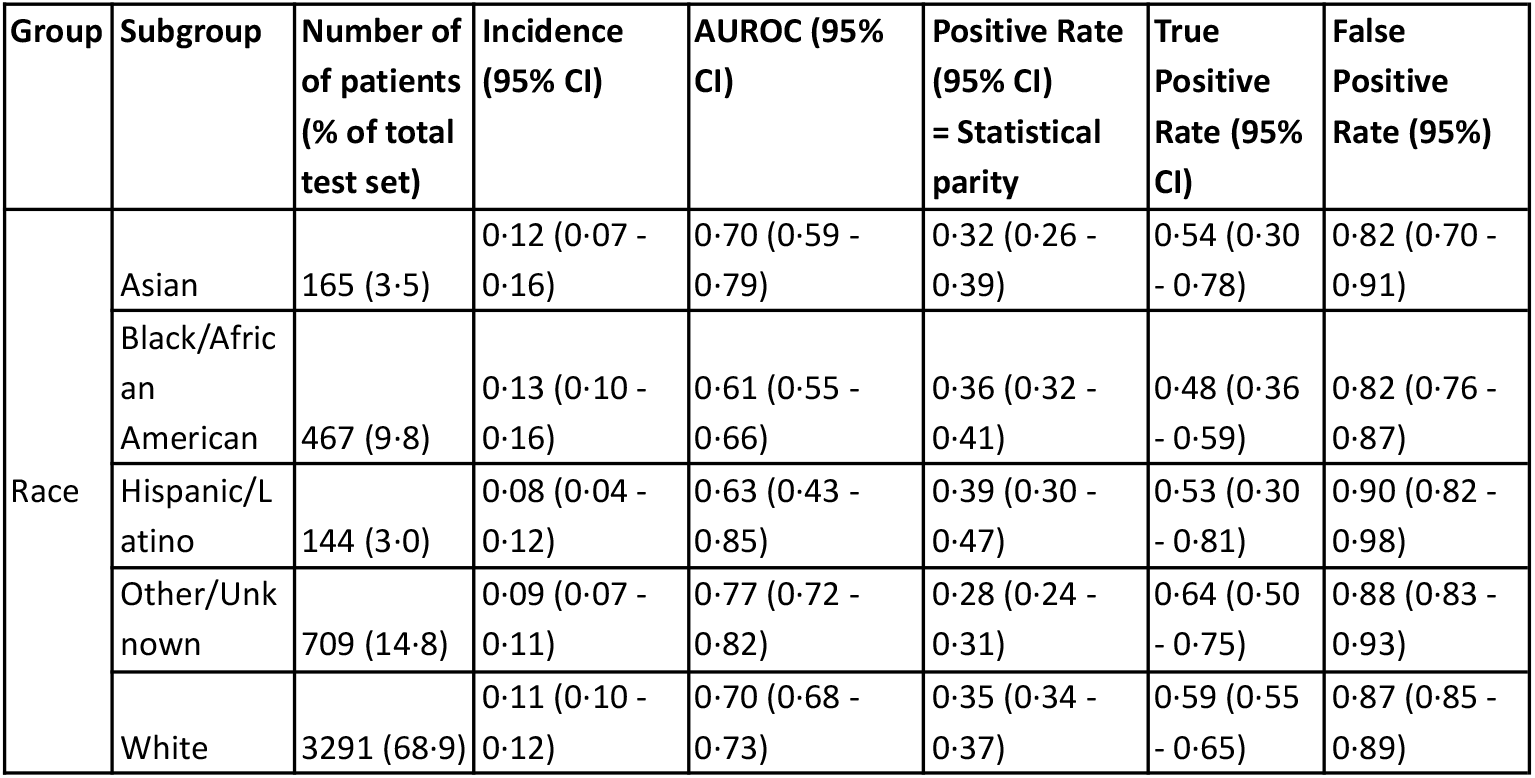
OUD (Opioid Use Disorder) resource allocation on MIMIC-IV test dataset prevalence and performance metrics without race included as predictive parameter. Metrics are calculated using a classification cut-off of 0‐1. To adhere to the notion of statistical parity, statistical parity should be equal across groups.

| Group | Subgroup | Number of<br>of patients<br>(% of total<br>test set) | Incidence<br>(95% CI) | AUROC (95%<br>CI) | Positive Rate<br>(95% CI)<br>= Statistical<br>parity | True<br>Positive<br>Rate (95%<br>CI) | False<br>Positive<br>Rate (95%) |
| --- | --- | --- | --- | --- | --- | --- | --- |
| Race | Asian | 165 (3.5) | 0.12 (0.07 - 0.16) | 0.70 (0.59 - 0.79) | 0.32 (0.26 - 0.39) | 0.54 (0.30 - 0.78) | 0.82 (0.70 - 0.91) |
|  | Black/African American | 467 (9.8) | 0.13 (0.10 - 0.16) | 0.61 (0.55 - 0.66) | 0.36 (0.32 - 0.41) | 0.48 (0.36 - 0.59) | 0.82 (0.76 - 0.87) |
|  | Hispanic/Latino | 144 (3.0) | 0.08 (0.04 - 0.12) | 0.63 (0.43 - 0.85) | 0.39 (0.30 - 0.47) | 0.53 (0.30 - 0.81) | 0.90 (0.82 - 0.98) |
|  | Other/Unknown | 709 (14.8) | 0.09 (0.07 - 0.11) | 0.77 (0.72 - 0.82) | 0.28 (0.24 - 0.31) | 0.64 (0.50 - 0.75) | 0.88 (0.83 - 0.93) |
|  | White | 3291 (68.9) | 0.11 (0.10 - 0.12) | 0.70 (0.68 - 0.73) | 0.35 (0.34 - 0.37) | 0.59 (0.55 - 0.65) | 0.87 (0.85 - 0.89) |

**Figure 4.**
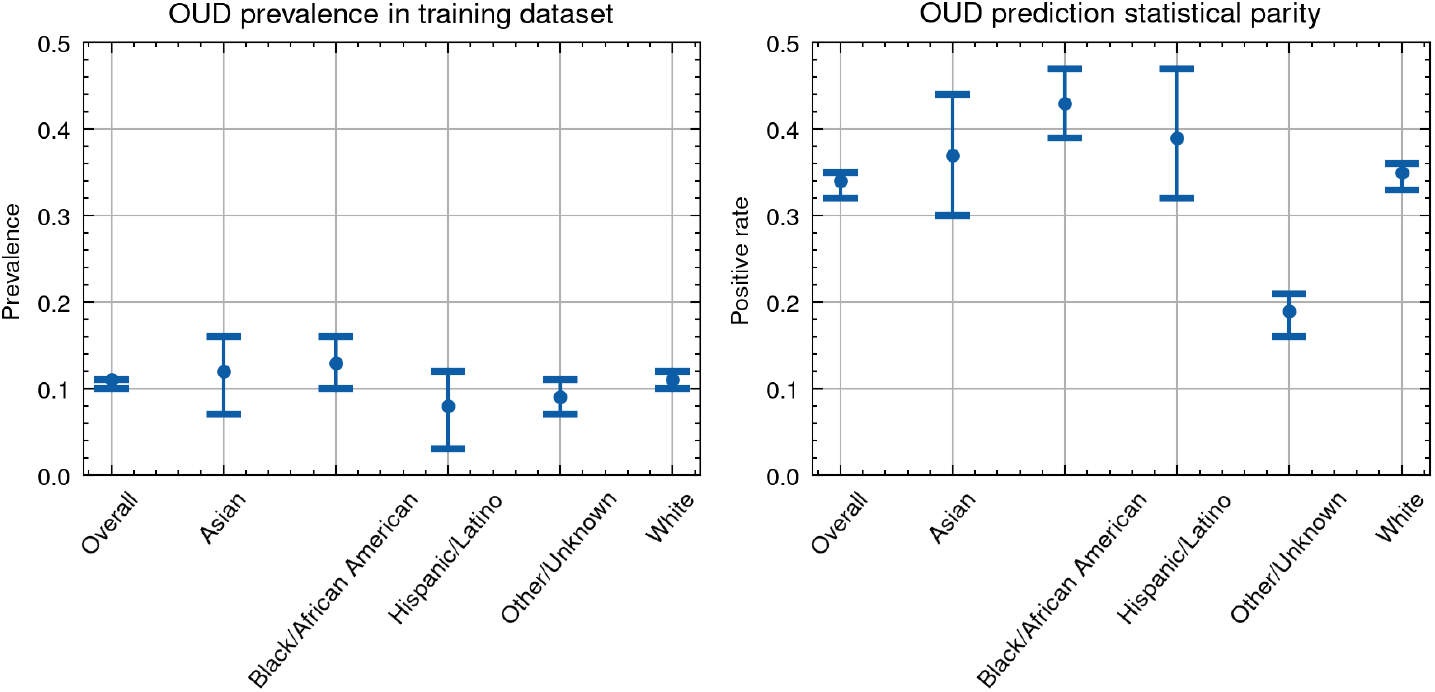
Baseline OUD rates (left) and statistical parity in terms of true positive rates (right) for opioid use disorder prediction in race subgroups

**Figure 5.**
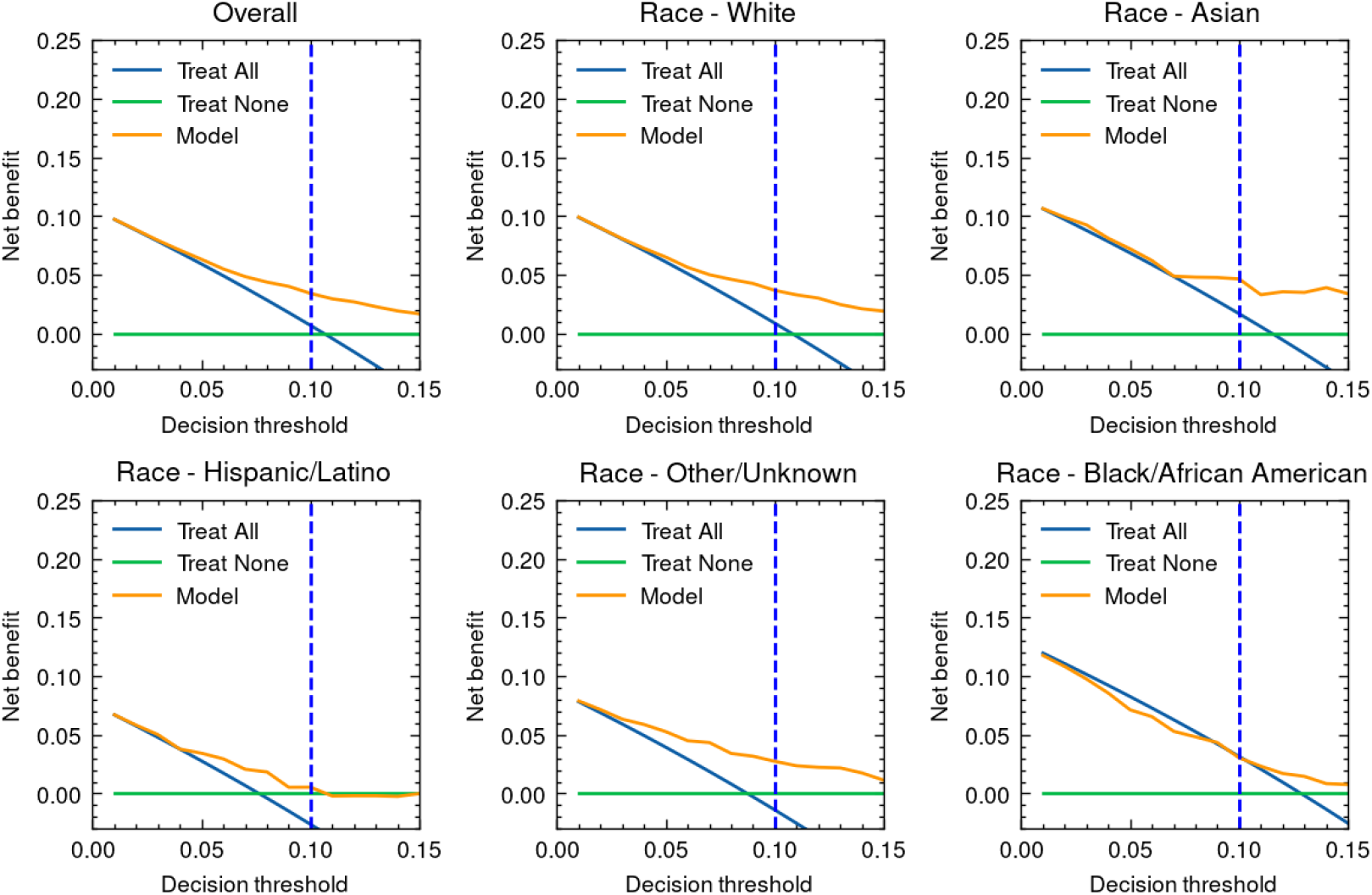
Clinical utility by means of net benefit curves for opoïd use prediction per race category. Classification cut-off was 0.10, meaning that patients with a risk higher than 10% will get the intervention.

## 6. Discussion

AI-based prediction models are moving from research to wider deployment, but biases in training data pose a challenge by potentially worsening healthcare disparities. Despite efforts to develop fair AI, there remains a lack of practical steps for evaluating fairness in clinical prediction models. This is partly due to the absence of a universally accepted definition of fairness, as seen in the 27 different metrics identified in recent literature. We propose a simplified framework for selecting relevant group-based fairness notions based on the model’s intended use, whether for decision support or informing with risk scoring, and the type of decision being influenced. This framework can assist AI model developers and assessors. While our focus is on group-based fairness, individual fairness may also need consideration depending on the clinical and ethical context.

Choosing and prioritizing fairness notions for AI-based prediction models is challenging due to the abundance of metrics and their incompatibility. Bias mitigation may improve fairness, but it is crucial to quantify fairness-performance trade-offs and assess their clinical impact [14, 31]. Some fairness measures are constrained by their dependence on epidemiological and statistical concepts, which may not hold across groups. Additionally, certain metrics involve causal claims, extending beyond the association-focused nature of AI models. We propose evaluating fairness and bias mitigation efforts based on clinical utility, calibration, performance metrics like AUROC, and/or statistical parity.

Removing variables like race is a common fairness strategy [19], but may be counterproductive if race is linked to outcomes [32]. Adjusting classification thresholds for statistical parity or equalized odds [33] also poses challenges, including implementation complexity and clinician bias. Mixed-race patients further complicate threshold application at the point of care. The most effective approach is ensuring unbiased, representative training data, though given limitations in observational data, other bias mitigation strategies are often required [25].

Quantitatively assessing fairness poses several challenges. We emphasize group-based fairness notions, whereas others state that fairness is “a latent construct imperfectly operationalised by statistical fairness measures” [34]. Furthermore, most quantitative measures rely on labels possibly affected by bias [24]. In response to these challenges, we advocate for an approach that incorporates quantitative and ethical fairness evaluations throughout the model development and validation lifecycle [35]. Our use cases show that visualizing fairness metrics supports ethical decision-making, ensuring a more comprehensive approach to fair AI systems.

This study has several limitations. Firstly, we focused on group-based notions of fairness and less on individual notions of fairness. While individual notions of fairness are important from a patient perspective, there are limited tools and metrics for systematically quantifying individual-level fairness, highlighting the need for more research on this topic. For example, it is challenging to determine similarity metrics for patients with good interpretability. Second, we focus on prediction models in terms of decision support and risk scores, with methods that may not apply to other applications, such as large language models. Since these types of AI models produce different kinds of outputs, their fairness requires further research.

In conclusion, we present a structured approach for evaluating fairness in AI-based prediction models, aiming to enable more equitable clinical AI implementations. Given the diverse and sometimes conflicting definitions of fairness, our proposed framework provides guidance for appropriate group-based fairness metrics to assess. Recognising the limitations of purely quantitative assessments, we recommend to, depending on the use case, assess clinical utility, calibration, performance-based measures, and/or statistical parity. The selection of these metrics should align with the specific distributive justice goal, whether to ensure equal benefit, performance, or resource allocation across patient groups. By addressing fairness through this comprehensive and multidimensional lens, our approach offers a practical foundation for developers and assessors, facilitating the transition from theoretical concepts of fairness to their practical application in AI-based healthcare systems.

## Data Availability

The MIMIC dataset used and/or analysed during the current study is available from the MIMIC repository. The LUMC dataset used and/or analysed during the current study is not publicly available as patients did not give informed consent for making their data public. The underlying code for this study is not publicly available but may be made available to qualified researchers upon reasonable request to the corresponding author.

## Author contributions

SM, YW, MA, BG, EW and TH conceived and designed the study. SM and MA performed the literature search. SM and YW acquired and varified the data and performed the analysis. SM, YW, MA, BG, EW and TH interpreted the results. SM created the figures. SM drafted the manuscript. All authors reviewed the data and revised the manuscript.

## Acknowledgements

Research reported in this publication was supported by the National Library Of Medicine of the National Institutes of Health under Award Number R01LM013362. The content is solely the responsibility of the authors and does not necessarily represent the official views of the National Institutes of Health.

## Data sharing statement

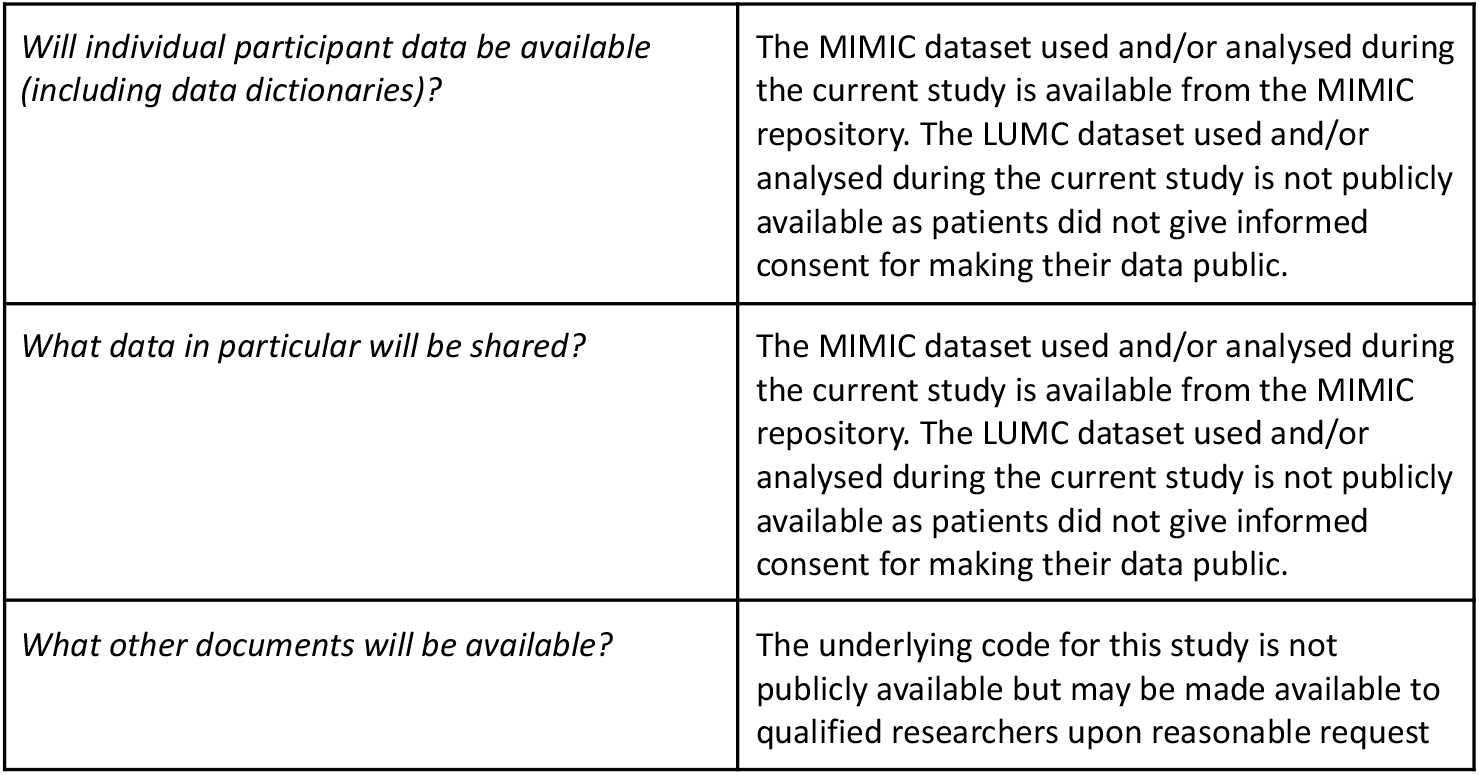
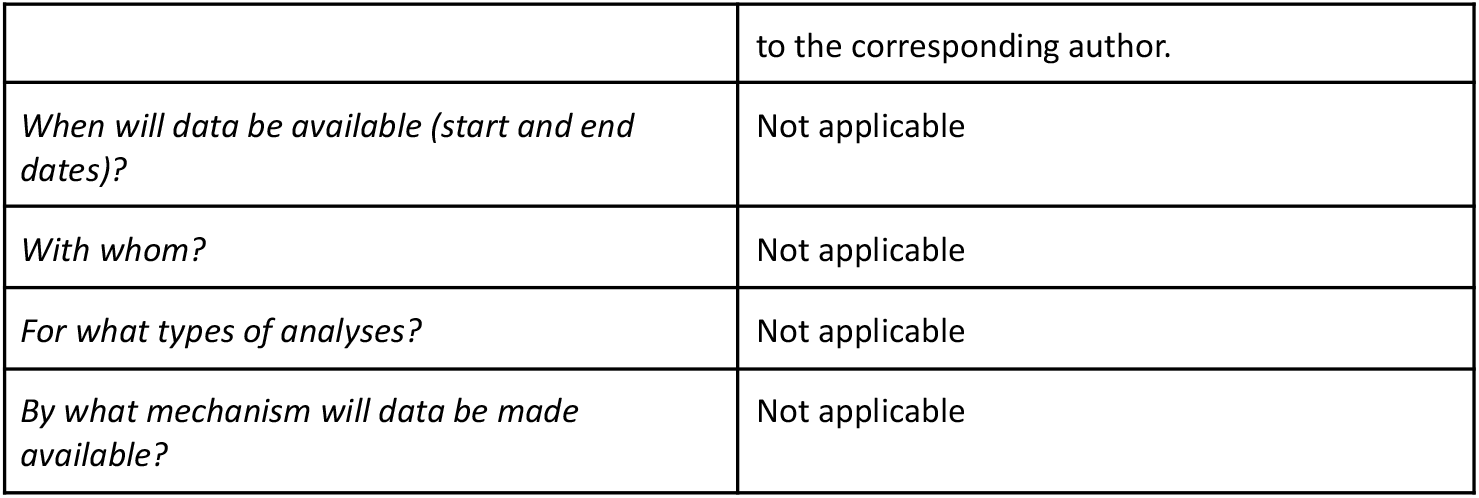

