## Supplementary Materials for "Navigating Fairness in AI-based Prediction Models: Theoretical Constructs and Practical Applications"

[1] Department of Intensive Care Medicine, The Leiden University Medical Center, Leiden, The Netherlands
[2] Healthplus.ai B.V.
[3] Department of Medicine (Biomedical Informatics), Stanford University, Stanford, CA, USA
[4] Department of Biomedical Datasciences, Leiden University Medical Center, Leiden, The Netherlands
[5] Department of Biomedical Data Science, Stanford University, Stanford, CA, USA

#### *Supplementary Materials*

#### Table of contents

**1. Bias mitigations and fairness evaluation throughout the model development lifecycle 2**
**2. Literature search 3**
**3. Fairness and bias definitions 5**
**4. Fairness evaluation metrics 8**
**5. Additional use case results 12**
**6. References 14**

### 1. Bias mitigations and fairness evaluation throughout the model development lifecycle

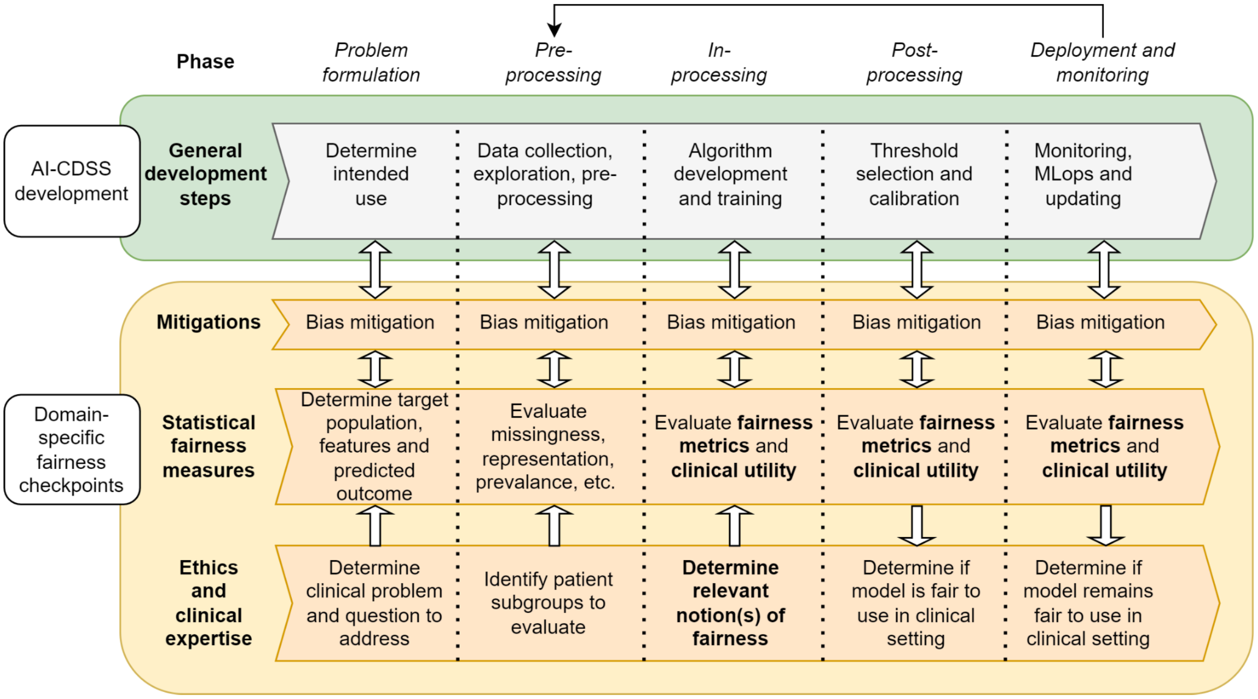


Figure S1: Developing fair AI-based clinical decision support systems throughout each step of the model development lifecycle. In the green frame, the general model development steps are outlined. In the yellow frame, the different domains of fairness evaluation and bias mitigation are outlined, distinguishing between ethical and clinical expertise, statistical fairness measures, and bias mitigation measures that interact with the general model development steps. In bold, the focus of the current work is indicated. AI-CDSS = AI-based clinical decision support systems, MLops = Integrating development and operations (DevOps) for efficient machine learning lifecycle management.

#### 2. Literature search

A systematic literature search was performed in PubMed and Web of Science to identify relevant review, survey and perspective papers on fairness evaluation of artificial intelligence models. The search was executed from inception to 18 January 2024, resulting in 231 unique references of which 184 were from PubMed and 76 (47 unique) from Web of Science. The following search query was used.

**(**(("Fairness"[ti] OR "Bias"[majr:noexp] OR "artificial intelligence bias"[title:~6] OR "AI bias"[title:~6] OR "machine learning bias"[title:~6] OR "deep learning bias"[title:~6] OR "algorithm bias"[title:~6] OR "algorithms bias"[title:~6]) AND ("Algorithms"[mesh] OR "AI"[ti] OR "Artificial Intelligence"[mesh:noexp] OR "Machine Learning"[mesh] OR "Deep Learning"[mesh] OR "artificial intelligence"[tw] OR "machine learning"[tw] OR "deep learning"[tw] OR "algorithm"[tw] OR "algorithms"[tw] OR "algorithm*"[tw]) AND ("review"[pt] OR "review"[ti] OR "overview"[ti] OR "synthesis"[ti] OR "metasynthesis"[ti] OR "survey"[ti] OR "systematic review"[pt] OR "systematic"[sb] OR "meta-analysis"[pt] OR "meta analysis"[tw] OR "metaanalysis"[tw] OR "meta analy*"[tw] OR "metaanaly*"[tw]) NOT ("risks of bias"[ti] OR "risks of bias"[ti])) **OR** (("Fairness"[ti] OR "Bias"[majr:noexp] OR "Bias"[ti] OR "artificial intelligence bias"[title:~6] OR "AI bias"[title:~6] OR "machine learning bias"[title:~6] OR "deep learning bias"[title:~6] OR "algorithm bias"[title:~6] OR "algorithms bias"[title:~6]) AND ("Algorithms"[mesh] OR "AI"[ti] OR "Artificial Intelligence"[mesh:noexp] OR "Machine Learning"[mesh] OR "Deep Learning"[mesh] OR "artificial intelligence"[tw] OR "machine learning"[tw] OR "deep learning"[tw] OR "algorithm"[tw] OR "algorithms"[tw] OR "algorithm*"[tw]) AND ("Critical Care"[Mesh] OR "Critical Care"[tw] OR "Intensive Care"[tw]) NOT ("risks of bias"[ti] OR "risks of bias"[ti])) **OR** (("Fairness"[tw] OR "Bias"[mesh:noexp] OR "artificial intelligence bias"[title/abstract:~6] OR "AI bias"[title/abstract:~6] OR "machine learning bias"[title/abstract:~6] OR "deep learning bias"[title/abstract:~6] OR "algorithm bias"[title/abstract:~6] OR "algorithms bias"[title/abstract:~6]) AND ("Algorithms"[mesh] OR "AI"[ti] OR "Artificial Intelligence"[mesh:noexp] OR "Machine Learning"[mesh] OR "Deep Learning"[mesh] OR "artificial intelligence"[tw] OR "machine learning"[tw] OR "deep learning"[tw] OR "algorithm"[tw] OR "algorithms"[tw] OR "algorithm*"[tw]) AND ("Terminology as Topic"[Mesh] OR "terminology"[tw] OR "terminologies"[tw] OR "nomenclature"[tw] OR "nomenclatur*"[tw] OR "ontology"[tw] OR "ontologies"[tw] OR "ontolog*"[tw] OR "definition"[tw] OR "definitions"[tw] OR concept*[tw]) AND ("review"[pt] OR "review"[ti] OR "overview"[ti] OR "synthesis"[ti] OR "metasynthesis"[ti] OR "survey"[ti] OR "systematic review"[pt] OR "systematic"[sb] OR "meta-analysis"[pt] OR "meta analysis"[tw] OR "metaanalysis"[tw] OR "meta analy*"[tw] OR "metaanaly*"[tw])) **OR** (("Fairness"[tw] OR "Bias"[mesh:noexp] OR "artificial intelligence bias"[title/abstract:~6] OR "AI bias"[title/abstract:~6] OR "machine learning bias"[title/abstract:~6] OR "deep learning bias"[title/abstract:~6] OR "algorithm bias"[title/abstract:~6] OR "algorithms bias"[title/abstract:~6]) AND ("Algorithms"[majr] OR "AI"[ti] OR "Artificial Intelligence"[majr:noexp] OR "Machine Learning"[majr] OR "Deep Learning"[majr] OR "artificial intelligence"[ti] OR "machine learning"[ti] OR "deep learning"[ti] OR "algorithm"[ti] OR "algorithms"[ti] OR "algorithm*"[ti]) AND ("prediction"[ti] OR "predict*"[ti] OR "decision support"[ti] OR "decision support"[ti] OR "Decision Support Systems, Clinical"[majr] OR "Decision Support Techniques"[majr] OR "Decision Support Systems, Management"[majr] OR "Clinical Decision-Making"[majr] OR "Decision Making"[majr] OR "decision"[ti] OR "decisions"[ti]) AND ("review"[pt] OR "review"[ti] OR "overview"[ti] OR "synthesis"[ti] OR "metasynthesis"[ti] OR "survey"[ti] OR "systematic review"[pt] OR "systematic"[sb] OR "meta-analysis"[pt] OR "meta analysis"[tw] OR "metaanalysis"[tw] OR "meta analy*"[tw] OR "metaanaly*"[tw]))**)**

Inclusion criteria were: 1) reviews, surveys, and perspectives on fairness and bias detection and mitigation for AI systems, 2) studies on fairness in either the clinical, healthcare, and general setting. Exclusion criteria were: 1) Studies focusing solely on one specific type of fairness and bias, 2) Studies focusing on fairness and bias specifically in other domains than healthcare.

After exclusion on title and abstract, 23 articles remained for full-text review. A total of 18 studies were included of which 16 originated from the systematic search and two additional studies were included from the references of other studies. See Table S1 for an overview of the included studies.

Table S1: Overview of included studies for literature review on fairness evaluation methods

| Study | Title | Type of study |
|---|---|---|
| Ueda 2024 | Fairness of artificial intelligence in healthcare: review and recommendations | Review |
| Abràmoff 2023 | Considerations for addressing bias in artificial intelligence for health equity | Perspective |
| Alves 2023 | Survey on fairness notions and related tensions* | Survey |
| Chen 2023 | Algorithmic fairness in artificial intelligence for medicine and healthcare | Perspective |
| Gray 2023 | Measurement and Mitigation of Bias in Artificial Intelligence: A Narrative Literature Review for Regulatory Science | Review |
| Liu 2023 | A translational perspective towards clinical AI fairness | Perspective |
| Pessach 2023 | A Review on Fairness in Machine Learning | Survey |
| Nazer 2023 | Bias in artificial intelligence algorithms and recommendations for mitigation | Review |
| Tang 2023 | What-is and How-to for Fairness in Machine Learning: A Survey, Reflection, and Perspective | Survey |
| Holm 2022 | Handle with care: Assessing performance measures of medical AI for shared clinical decision-making | Review |
| Varona 2022 | Discrimination, Bias, Fairness, and Trustworthy AI | Review |
| Xu 2022 | Algorithmic fairness in computational medicine | Review |
| Fletcher 2021 | Addressing Fairness, Bias, and Appropriate Use of Artificial Intelligence and Machine Learning in Global Health | Review |
| Makhlouf 2021 | Machine learning fairness notions: Bridging the gap with real-world applications | Survey |
| Mehrabi 2021 | A Survey on Bias and Fairness in Machine Learning | Survey |
| Caton 2020 | Fairness in Machine Learning: A Survey | Survey |
| Paulus 2020 | Predictably unequal: understanding and addressing concerns that algorithmic clinical prediction may increase health disparities | Perspective |
| Rajkomar 2018 | Ensuring Fairness in Machine Learning to Advance Health Equity | Perspective |

#### 3. Fairness and bias definitions

Table S2: Definitions of fairness and bias used in included studies

| Study | Definition of bias | Definition of fairness |
|---|---|---|
| Ueda 2024 | N/A | Fairness in healthcare is a multidimensional concept that includes the equitable distribution of resources, opportunities, and outcomes among diverse patient populations |
| Abràmoff 2023 | Bias can be quantified as differential impact of a healthcare process on a particular group. | N/A |
| Alves 2023 | N/A | Unfair decisions: typically discriminating against disadvantaged populations such as racial minorities, women, poverty-stricken districts etc. |
| Chen 2023 | N/A | A concept for defining, quantifying and mitigating unfairness from machine-learning predictions that may cause disproportionate harm to individuals or groups of individuals. Fairness is a formalization of the minimization of disparate treatment and impact. There are multiple criteria for the quantification of fairness, yet they typically involve the evaluation of differences in performance metrics. |
| Gray 2023 | Oxford English Dictionary: "a tendency, inclination or leaning towards a particular characteristic, behavior, etc." | N/A |
| Liu 2023 | Difference is not equal to bias. | A fair model is expected to perform equally across subgroups defined by sensitive variables. Fairness is absence of bias: “Bias and fairness are two concepts that usually oppose each other: a decision is unfair if it is biased towards (or against) any individual or subpopulation" |
| Pessach 2023 | We use the terms bias, discrimination, and unfairness interchangeably with similar meanings, as is commonly done in the algorithmic fairness literature | We use the terms bias, discrimination, and unfairness interchangeably with similar meanings, as is commonly done in the algorithmic fairness literature |
| Nazer 2023 | N/A | N/A |

| Study | Definition of bias | Definition of fairness |
| --- | --- | --- |
| Tang 2023 | N/A | Machine learning literature has proposed a deluge of algorithmic fairness definitions, each of which comes with explicit or implicit assumptions on the discrimination of interest and the corresponding mathematical formulation that captures it. |
| Holm 2022 | N/A | The fairness of an algorithm in terms of equality across groups in one or more performance measures (classification parity definition of fairness) |
| Varona 2022 | Bias represents the action, while discrimination manifests itself in the result of using certain attributes in the decision-making process. | Gives several definitions from literature:<br>Technical point of view: the actions performed to optimize search engines or ranking services without altering or manipulating them for purposes unrelated to the users' interest.<br>Ability to treat all similar individuals or groups equally and the AI system's inability to produce harm in any possible way. |
| Xu 2022 | Defines different types of computational biases. | N/A |
| Fletcher 2021 | A systematic tendency in a model to favor one demographic group vs another, which can be mitigated<br>but can lead to unfairness. Bias is independent of ethics. | Involves examining the impact on various demographic groups and choosing one of several mathematical definitions of group fairness that will adequately satisfy the desired set of legal, cultural, and ethical requirements |
| Makhlouf 2021 | N/A | Fairness in machine learning can be categorized according to two dimensions, namely, the task and the type of learning. |
| Mehrabi 2021 | Bias can be considered as a source for unfairness that is due to the data collection, sampling, and measurement. | Fairness is the absence of any prejudice or favoritism toward an individual or group based on their inherent or acquired characteristics |
| Caton 2020. | N/A | No general consensus as to what constitutes fairness or if it can/should be quantified |
| Paulus 2020 | We use the term algorithmic bias (in distinction to fairness) specifically to refer to these issues related to model design, data and sampling that may | Not one definition |

| Study | Definition of bias | Definition of fairness |
|---|---|---|
| | disproportionately affect model performance in a certain subgroup. | |
| Rajkomar 2018 | Gives definitions for different types of biases. | Is not just about preventing a model from harming a protected group; it may also help focus care where it is really needed. |

### 4. Fairness evaluation metrics

Table S3: Identified fairness metrics, alternative names, and description

| Scope of application | Fairness metric name | Alternative names | Description | References |
|---|---|---|---|---|
| 1: Group | Equalized odds | Separation, conditional procedure accuracy equality, disparate mistreatment, error rate balance | Different groups deal with similar odds. **True positive rate and false positive rate** are equal among groups. | Xu et al. Hardt et al. Mehrabi et al. Alves et al. Chen et al. Gray et al. Holm et al. Tang et al. Liu et al. Makhlouf et al. Paulus et al. Pessach et al. Rajkomar et al. Caton et al. |
| 1: Group | Statistical parity | Demographic parity, independence, equal acceptance rate, benchmarking, group fairness, equal allocation, equal outcomes | Requires that the **proportion of positive/negative predicted cases** is the same across groups and the total population. | Xu et al. Calders et al. Mehrabi et al. Alves et al. Chen et al. Gray et al. Tang et al. Liu et al. Makhlouf et al. Paulus et al. Pessach. Rajkomar et al. Caton et al. Fletcher et al. |
| 1: Group | Conditional statistical parity | Conditional discrimination-aware classification | Like statistical parity, but then controlling on a **set of conditioning features**, that are correlated with the sensitive feature. | Alves et al. Liu et al. Makhlouf et al. |
| 1: Group | Overall accuracy equality | | **Accuracies are equal across groups.** | Alves et al. Liu et al. Makhlouf et al. |
| 1: Group | Equal opportunity | False negative error balance rate | **True positive rates are equal across groups.** | Xu et al. Hardt et al. Mehrabi et al. Alves et al. Liu et al. Makhlouf et al. Pessach et al. Rajkomar et al. Caton et al. Fletcher et al. |
| 1: Group | Predictive equality | False positive error balance | **False positive rates are equal across groups.** | Alves et al. Liu et al. |
| 1: Group | Predictive parity | Outcome test | **Positive predictive values are equal across groups.** | Alves et al. Chen et al. Gray et al. Liu et al. Makhlouf et al. Paulus et al. Rajkomar et al. |
| 1: Group | Conditional use accuracy | Sufficiency, equal predictive value | **Positive predictive values and negative predictive values are equal across groups.** | Alves et al. Holm et |

| Scope of application | Fairness metric name | Alternative names | Description | References |
|---|---|---|---|---|
| | equality | | | al. Liu et al. Makhlouf et al. Caton et al. |
| 1: Group | xAUC | Disparity in bipartite-ranking metrics | The xAUC (cross area under the curve) is a metric to assess the disparate impact of risk scores and quantifies the difference in the likelihood of ranking a randomly chosen positive example from one protected group higher than a negative example from another group, and vice versa. | Liu et al. |
| 1: Group | Test fairness | Calibration, matching conditional frequencies, well calibration | Calibration is equally good in among groups: "that for any predicted probability score S, people in both protected and unprotected groups must have equal probability of correctly belonging to the positive class." | Mehrabi et al. Alves et al. Makhlouf et al. Paulus et al. Caton et al. Fletcher et al. |
| 1: Group | Balance | Positive balance, negative balance | Balance for positive class and balance for negative class. Assesses predicted probabilities in both classes. | Alves et al. Makhlouf. Paulus et al. Caton et al. |
| 1: Group | Total fairness | | Statistical parity, equalized odds, conditional use accuracy equality, and treatment equality are satisfied. | Alves et al. Makhlouf |
| 1: Group | Negative predictive parity | | **Negative predictive values are equal across groups.** | Makhlouf et al. Paulus et al. |
| 1: Group | Fairness on average causal effect | | Matches individuals with different protected attributes which are closest concerning a distance measure, and calculates difference in the outcome (**counterfactual approach**) | Tang et al. |
| 1: Group | Equality of effort | | The equality of effort notation helps answer questions like to what extent a legitimate variable should change to make a particular individual achieve a certain outcome level and address the concerns about whether the efforts made to achieve the same outcome level for individuals from the protected group and that from the unprotected group are different. (**counterfactual**) | Tang et al. |
| 1: Group | No direct/indirect discrimination | | No Direct Discrimination (based on path-specific causal effects from the protected feature to the predictor) and No Indirect Discrimination (considering paths involving redlining attributes). | Tang et al. |
| 1: Group | Treatment equality | | **False negatives and false positives are equal across groups.** | Mehrabi et al. Alves et al. |
| 1: Group | Bayesian fairness | | Considers scenarios (different plausible model parameter estimates) in the resulting | Caton et al. |

| Scope of application | Fairness metric name | Alternative names | Description | References |
|---|---|---|---|---|
| | | | fairness/unfairness. Extended concept of balance. | |
| 2: Individual | Counterfactual measures | Counterfactual fairness, causal discrimination | If for two similar patients, only the protected attribute/sensitive variable is changed, the predicted outcome should not change. Predictions remain the same in both the actual world and a hypothetical world where an individual's attributes, such as race or gender, are different. Causal models are used to address potential biases in the data. | Xu et al. Kusner et al. Mehrabi et al. Alves et al. Gray et al. Liu et al. Makhlouf et al. Caton et al. |
| 2: Individual | Fairness through awareness | Individual fairness | Similar individuals have similar predictions, according to similarity metrics. | Xu et al. Dwork et al. Mehrabi et al. Alves et al. Liu et al. Pessach et al. |
| 2: Individual | Generalized Entropy Index | | Accounts for variations in an individual's prediction accuracy compared to the average prediction accuracy across a group. | Caton et al. |
| 2: Individual | No unresolved discrimination | No proxy discrimination | Through a conditioning variable, e.g., education level in women, the predicted outcome may be influenced, but there is no direct link between the sensitive variable and the predicted outcome. | Tang et al. Makhlouf et al. Mehrabi et al. |
| 2: Individual | Fairness through unawareness | Blindness, unawareness, anti-classification, individual fairness | No protected attributes used in decision-making process. | Xu et al. Kusner et al. Mehrabi et al. Alves et al. Tang et al. Liu et al. |
| 2: Individual | Individual fairness in hindsight | | Extends the concept of individual fairness to account for temporal considerations. | Tang et al. |
| 3: Subgroup | Subgroup fairness | | Subgroup fairness aims to combine the strengths of both group and individual fairness concepts. While distinct from these notions, it leverages them to achieve improved outcomes. It applies a group fairness constraint, such as equalizing false positive rates, and evaluates whether this constraint is met across a wide range of subgroups. | Mehrabi et al. |
| 4: Distributive | Variance (standard deviation) of the quantities | | Variance or standardized deviation of the quantities (e.g., accuracy, loss, etc), emphasizing the equality of quantities received by participants. | Liu et al. |

| Scope of application | Fairness metric name | Alternative names | Description | References |
| --- | --- | --- | --- | --- |
| 4: Distributive | Reward based on correlation | | The correlation between party contributions (combining standalone model accuracies and sharing levels) and party rewards (final model accuracies). The correlation coefficient (rxy) is calculated to assess fairness, with a higher value indicating good fairness (within [-1,1]), while a negative coefficient implies poor fairness. | Liu et al. |

#### 5. Additional use case results

Table S4: PERISCOPE prevalence and performance metrics

| Group | Subgroup | Number of procedures (% of total testset) | Prevalence (95% CI) | AUROC (95% CI) | Calibration slope (95% CI) | Calibration intercept (95% CI) |
|---|---|---|---|---|---|---|
| Gender | Female | 4826 (53.5) | 0.11 (0.10-0.11) | 0.80 (0.78-0.81) | 0.81 (0.75-0.88) | -0.08 (-0.16-0.01) |
| | Male | 4197 (46.5) | 0.16 (0.15-0.17) | 0.82 (0.80-0.83) | 0.92 (0.83-1.00) | -0.23 (-0.30--0.12) |

Table S5: OUD (OipoÏd Use Disorder) resource allocation on MIMIC-IV test dataset prevalence and performance metrics when race is included as predictive parameter. Metrics are calculated using a classification cut-off of 0.1. To adhere to the notion of statistical parity, the positive rate should be equal across groups.

| Group | Subgroup | Number of of patients (% of total testset) | Prevalance (95% CI) | AUROC (95% CI) | Positive Rate (95% CI) = Statistical parity | True Positive Rate (95% CI) | False Positive Rate (95%) |
|---|---|---|---|---|---|---|---|
| Race | Asian | 165 (3.5) | 0.12 (0.07 - 0.16) | 0.75 (0.64 - 0.87) | 0.37 (0.32 - 0.42) | 0.66 (0.44 - 0.84) | 0.88 (0.78 - 0.96) |
| | Black/ African American | 467 (9.8) | 0.13 (0.10 - 0.16) | 0.61 (0.54 - 0.68) | 0.43 (0.39 - 0.46) | 0.53 (0.40 - 0.64) | 0.86 (0.81 - 0.91) |
| | Hispanic/ Latino | 144 (3.0) | 0.08 (0.04 - 0.12) | 0.67 (0.47 - 0.83) | 0.38 (0.32 - 0.47) | 0.53 (0.27 - 0.81) | 0.91 (0.83 - 0.97) |
| | Other/ Unknown | 709 (14.8) | 0.09 (0.07 - 0.11) | 0.77 (0.73 - 0.82) | 0.19 (0.16 - 0.22) | 0.48 (0.35 - 0.62) | 0.76 (0.70 - 0.83) |
| | White | 3291 (68.9) | 0.11 (0.10 - 0.12) | 0.71 (0.69 - 0.74) | 0.35 (0.34 - 0.36) | 0.62 (0.57 - 0.68) | 0.87 (0.86 - 0.89) |

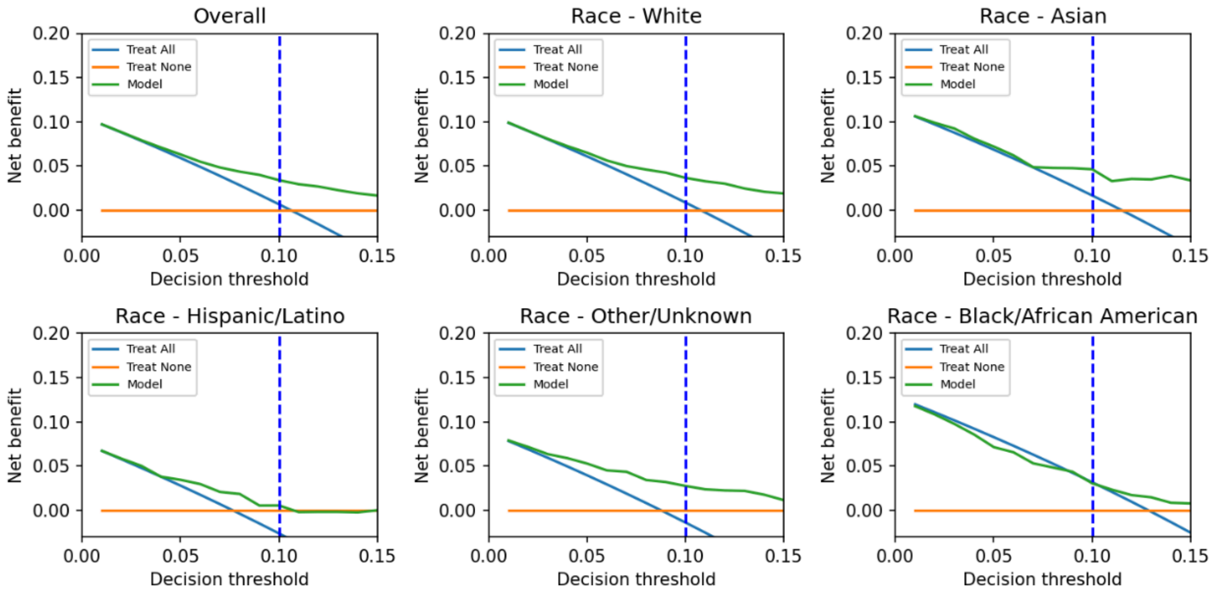


Figure S2: Net benefit curves for OUD prediction when race is included as predictor.

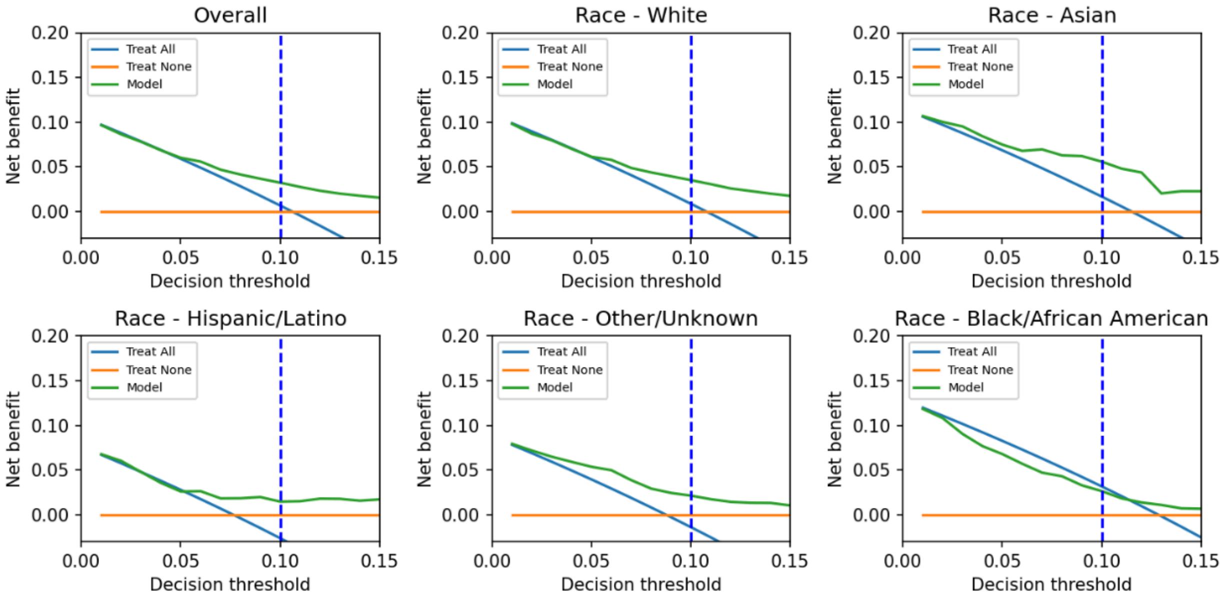


Figure S3: Net benefit curves after oversampling on the training dataset to obtain equally sized subgroups
